# Intracranial pressure-flow relationships in traumatic brain injury patients expose gaps in the tenets of models and pressure-oriented management

**DOI:** 10.1101/2024.01.17.24301445

**Authors:** JN Stroh, Brandon Foreman, Tellen D Bennett, Jennifer K Briggs, Soojin Park, David J Albers

## Abstract

**Background:** The protocols and therapeutic guidance established for treating traumatic brain injuries (TBI) in neurointensive care focus on managing cerebral blood flow (CBF) and brain tissue oxygenation based on pressure signals. The decision support process relies on assumed relationships between cerebral perfusion pressure (CPP) and blood flow, pressure-flow relationships (PFRs), and shares this framework of assumptions with mathematical intracranial hemodynamic models. These foundational assumptions are difficult to verify, and their violation can impact clinical decision-making and model validity.

**Method:** A hypothesis– and model-driven method for verifying and understanding the foundational intracranial hemodynamic PFRs is developed and applied to a novel multi-modality monitoring dataset.

**Results:** Model analysis of joint observations of CPP and CBF validates the standard PFR when autoregulatory processes are impaired as well as unmodelable cases dominated by autoregulation. However, it also identifies a dynamical regime-or behavior pattern-where the PFR assumptions are wrong in a precise, data-inferable way due to negative CPP-CBF coordination over long timescales. This regime is of both clinical and research interest: its dynamics are modelable under modified assumptions while its causal direction and mechanistic pathway remain unclear.

**Conclusions:** Motivated by the understanding of mathematical physiology, the validity of the standard PFR can be assessed *a)* directly by analyzing pressure reactivity and mean flow indices (PRx and Mx) or *b)* indirectly through the relationship between CBF and other clinical observables. This approach could potentially help personalize TBI care by considering intracranial pressure and CPP in relation to other data, particularly CBF. The analysis suggests a threshold using clinical indices of autoregulation jointly generalizes independently set indicators to assess CA functionality. These results support the use of increasingly data-rich environments to develop more robust hybrid physiological-machine learning models.

**Author Summary:** The current understanding of pressure-flow relationships used in neurocritical decision making are incomplete, and a novel dataset begins to illuminate what is missing.

## 1 Introduction

Clinical management is essential for improving patient neurological outcome following traumatic brain injury (TBI), an annual contributor to tens of thousands of fatalities in the USA (https://wonder.cdc.gov/mcd. html). TBI patients risk insults such as elevated intracranial pressure (ICP) and cerebral ischemia, compounding the prospect of secondary injuries such as hemorrhage and hypoxic tissue death. Contributors to these potential problems include impaired or overburdened cerebral autoregulation, an endogenous collection of perfusion control mechanisms reacting via changes in cerebral vessel diameter [9]. In TBI management [42], observable pressures, ABP and ICP, play a critical guiding role in intervention strategies. Meanwhile, blood transport, as quantified by cerebral perfusion and cerebral blood flow (CBF), is not typically measured and does not currently factor in care protocols. This work focuses on a core tenet of TBI management [4]: the relationship between commonly observed hemodynamic pressures and the associated, but typically unobserved, perfusion. TBI management protocols currently set thresholds for ICP or cerebral perfusion pressure (CPP, the mean ABP-ICP difference) to improve categorical clinical outcomes [7, 25]. The approach is contentious (*e.g.*, [2, 19]) and lacks personalization [43] despite improvements afforded by integrating *e.g.*, brain tissue oxygenation (P_bt_O_2_) [8], demographics, or disease severity [40]. Protocol objectives aim to maintain adequate perfusion for metabolic processes while minimizing the risk of secondary insults such as hyperemia and vascular barotrauma [18, 24]. Recent work [34] identified that optimized CPP is not always associated with optimal flow and oxygenation, further highlighting the insufficiency of pressure-targeted approaches. Directly or indirectly, pressure-guided protocols address perfusion, broadly referred to here as cerebral blood flow (CBF).

Despite both invasive[48, 37] and noninvasive [39, 51] collection methods, perfusion and CBF data are infrequently measured. CBF and observed pressures entwine with cerebral autoregulation (CA), pressure-responsive vasocontrol and other flow-regulatory mechanisms [9] that influence the pressure-flow relationship (PFR). The role of CA is implicit in the pressure reactivity index (PRx) [11, 10], a quantification of CA function from ABP and ICP. More explicitly, the mean flow index (Mx) [12] gauges CA via correlation of middle cerebral artery blood velocity (or its flow [3]) with CPP (or ABP, assuming ICP is constant). Existing studies associate favorable patient outcome with Mx or PRx lying below 0.3, establishing this value as a heuristic indicator of CA functionality [26, 40, 36].

Hemodynamic pressures (ABP, ICP), perfusion (CBF), and assemblage of CA processes form an inter-dependent system of intracranial hemodynamics (ICHD, Fig 1). In this system, Mx and PRx quantify specific aspects of ICHD interaction. A more complete description of ICHD includes cranial volume capacity, cranial blood volume, and additional extra-hemodynamic factors. Conceptually, this ICHD model (discussed below) asserts that perfusion is governed by CPP dynamics mitigated by CA processes.

**Figure 1:**
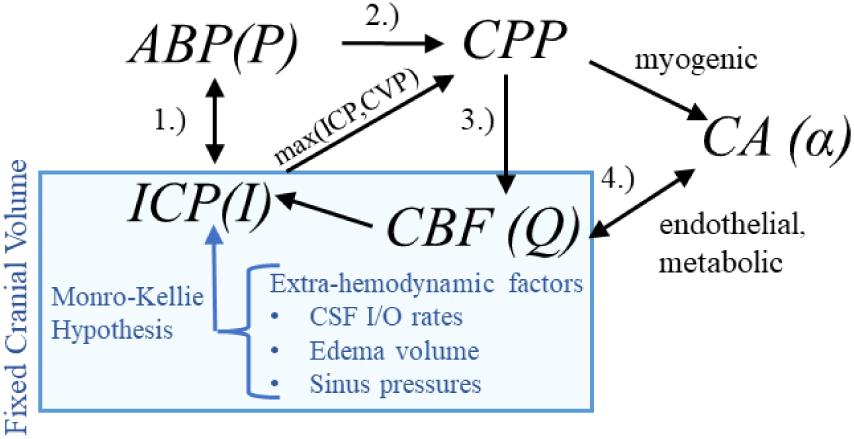
Hemodynamics relationships associated with ICP. In the first relationship (1.), ABP acts as the system inflow pressure with ICP opposing outflow pressure when it exceeds central venous pressure (CVP). In the second (2.), the ABP-ICP difference defines the background pressure gradient (CPP). The third (3.) is the pressure-flow relationship: CBF is determined by the pressure gradient subject under vasoregulation and other influences. The relationship (4.) identifies the co-dependence of functioning CA and CBF (or available oxygen/nutrients beyond the scope of this discussion). *CSF – cerebrospinal fluid*

### 1.1 Pressure-flow relationships

Defining both simulation models (*e.g.*,[23]) and clinical indices as PRx, Mx, and CPPopt [1, 45] requires assuming relationships among pressure, flow, and autoregulation. In elementary fluid settings, for example, increases in pressure difference imply increases in flow co-determined by changes in vessel diameter. The fluid relationship synthesized with knowledge of CA’s mitigating function motivates PFR hypotheses according to sign and potential causal direction:

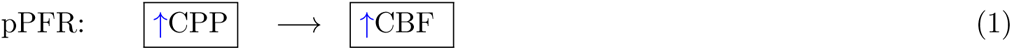

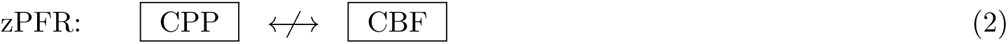

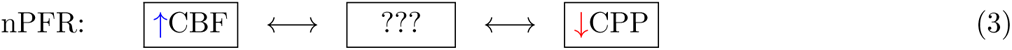

The positive PFR (pPFR, Eq (1)) posits that CBF results from pressure-driven processes regulated by variable vasoresistance. This pressure-passive situation is associated with autoregulatory impairment and serves as a baseline hypothesis in this work. The pPFR, for example, is assumed by cerebrovascular resistance (CVR, the mean ratio of CBF to CPP, *i.e.*, CBF = CPP/CVR [32, 35, 6]) as a relative measure of CA function. Under fixed CPP, flow increase is mathematically associated with decreased CVR and tied to CA-driven vasodilation. However, CVR is expected to remain positive so that CBF changes are positively or neutrally related CPP changes. Increased CBF under decreased CPP requires the vasodilation to be modeled by negative resistance.

The zero PFR (zPFR, Eq (2)) indicates decoupled CBF and CPP, presumably by functional autoregulation yielding no net correlation between flow and pressure. The negative PFR (nPFR, Eq (3)) is an alternate hypothesis comprising anti-correlated pressure-flow dynamics resulting from unspecified pathways. The dynamics of ICHD under zPFR and nPFR violate the positive pressure-flow association enforced by pPFR-based models, and these cases are therefore expected to be poorly modeled.

This study hypothesizes that functional CA of zPFR is an equilibrium between positive and negative PFRs over 2-hour timescales with neither being dominant. Namely, impaired CA corresponds to pPFR and functional CA to zPFR, while the nPFR is not expected to persist over such long timescales.

### 1.2 Purpose & Outline

This work examines the validity of assumptions made by conceptual hemodynamics models which relate observable pressures to the perfusion quantities affected by pressure-guided TBI management protocols. The lack of necessary ICHD data has prevented the validation at multi-hour timescales of the pressure-flow relationships that underlie pressure-oriented TBI therapies and autoregulatory measures. This work exploits a single-center multi-modal monitoring (MMM) observation of neurocritical patients to investigate model assumption consistency with observed signals in clinically relevant TBI cases. It assesses model sufficiency in application to clinically relevant cases. Its goals are to compute assumptions’ failure, to bring knowledge limits into focus, and to propose practicable model domain improvements.

The analysis of this work uses model-aided PFR categorization of clinical data to identify patterns of intracranial hemodynamics inaccurately represented by pPFR assumptions. Cases with persistent negative pressure-flow association (nPFR) compose a sizable portion of examined data; alternate assumptions not presently formulated are required to inform neurocritical care of patients presenting these dynamics. Empirical characteristics of CA indexes associated with pPFR and non-pPFR data indicate that pressure-flow relationships generalize qualitative assessment of autoregulatory function and suggests a pathway for including CBF data into the decision support process.

## 2 Method

This research explores PFRs by analyzing neurocritical patient data through model-simulated hemodynamics. These vital aspects are presented prior to defining the experiments and metrics used to assess PFRs. Simulations generate simulated observations whose appropriateness categorizes pressure-flow association of the data. This hypothesis-driven assumes that PFR timescales manifest between timescales of 1–2 minutes and 2 hours.

### 2.1 The Neurocritical Care Dataset

The University of Cincinnati obtained continuous multimodal monitoring (MMM) data from neurointensive TBI patients between 2014 and 2019 (Table 1). The collection occurred with prior consent under local institutional review board authorization (UCIRB #18-0743), These single-center data include concurrent records of ABP and ICP (125 Hz), plus brain tissue perfusion (1 Hz), brain tissue oxygenation (P_bt_O_2_, 1 Hz), and intracranial temperature (ICT, 0.5–2 Hz) observed by a Bowman probe (https://hemedex. com/products/bowman-perfusion-monitoring-system/). Further patient data include extra-ventricular drainage (EVD) transducer observations (which may be open or clamped), systemic monitors (EtCO_2_, central venous pressure, temperature), temperature management system use, and PRx computed bedside through Moberg CNS monitor systems (https://www.moberg.com/products/cns-monitor). The recording and storage process discretize patient data into epochs of irregular length that were analyzed separately. EVD observation of ICP was not used because data lacked clamp and calibration status timeseries needed for hour-scale analysis. Endnotes of this work address data access.

**Table 1:** Cohort description of University of Cincinnati MMM dataset. Data described as mean*±*standard deviation, median [quartile range], or proportion (%) as appropriate.

| Variable | Value (N=25) |
| --- | --- |
| **Demographics & Injury Characteristics** |  |
| Age (years) | 40.2±17.7 |
| Sex (male) | 20 (80%) |
| Injury mechanism |  |
| Motor Vehicle Collision | 12 (48%) |
| Fall | 10 (40%) |
| Other | 3 (12%) |
| $\Delta t$ Injury to Admission (hrs) | 1.5 [0.8–2.0] |
| **Admission Status** |  |
| Glasgow Coma Score (GCS) | 3 [3, 5] |
| Motor Subscore | 1 [1, 3] |
| Unreactive Pupil (1 or both) | 11 (44%) |
| Pre-hospital Hypoxia | 4 (16%) |
| Pre-hospital Hypotension | 0 (0%) |
| Rotterdam CT Score | 4 [3, 5] |
| Injury Severity Score | 25.7±9.5 |
| **Encounter Therapies** |  |
| Mechanical Ventilation | 25 (100%) |
| Decompressive Hemi-craniectomy | 9 (36%) |
| **Monitoring Data** |  |
| $\Delta t$ Injury to Moberg CNS data (hrs) | 13.1 [7.2, 17.7] |
| Duration of Moberg CNS data (hrs) | 32.1 [22.5, 57.3] |
| ICP Monitoring | 16 (64%) |
| $\Delta t$ Injury to ICP Monitoring (hrs) | 6.6 [5.5, 14.6] |
| ICP Monitor Type |  |
| Parenchymal Monitor | 11 (69% of 16) |
| External Ventricular Drainage Catheter | 0 (0%) |
| Both | 4 (25% of 16) |
| ICP Available Waveform Signal Data | 16 (100% of 16) |
| ICP Available Numeric Data | 16 (100% of 16) |
| PbtO<sub>2</sub> Monitoring | 16 (64%) |
| PbtO<sub>2</sub> Available Numeric Data | 16 (100% of 16) |
| **Outcome** |  |
| Hospital Length of Stay (days) | 12.8 [11.8, 20.1] |
| In-Hospital Mortality | 8 (32%) |
| Withdrawal of Life-Sustaining Therapy | 6 (75% of 8) |
| 6-Month Mortality | 10 (40%) |
| 6-Month Glasgow Outcome Scale-Extended | 3 [1, 6] |

### 2.2 Simulating Hemodynamics and Metrics

This study outlines a common conceptual ICHD framework used across various models to ensure generalizability. The system comprises ABP (*P*), ICP (*I*), CA processes (*α*), and CBF (*Q*) whose interactions appear in Fig 1. The model hypothesizes that blood flow results from the arteriovenous pressure gradient and interactive CA processes. More complex hypotheses considering cerebrospinal fluid [11] or volume distributions [38] are excluded; their associated models are difficult to identify and are too computationally expensive for hour-scale simulation [44].

The pressure gradient (*∇p*) is the assumed driving force of the system, and its average is the analogue of CPP. Ignoring spatial heterogeneity of vasculature, the zero-dimensional compartmental ICHD system reduces to:

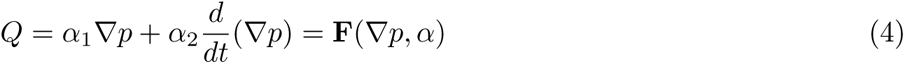

where components of the vector *α* = (*α*_1_*, α*_2_) parametrize CA effects on vessel bed conductance and compliance, respectively. Components of *α* are assumed to be non-negative due to assumption of pPFR (Eq (1)) except as transient behavior. The compartmental model Eq (4) is adopted in various models [23, 15, 22] or approximates those with more complex CA [46, 47], with spatially-distributed volume components [21, 49, 38], or with venous dynamics [41].

Temporal averaging produces the common CVR definition *α*_1_ = *∇p/Q* because the compliant storage term of Eq 4 is negligible in relation to the total flow. The time-averaging in differential form relates changes in CPP to changes in CBF, 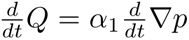, which is relevant to analysis through correlation. Importantly, it applies to nPFR dynamics with the inclusion of a negative sign while maintaining *α >* 0. *The relationship between this sign and PFR choice is exploited to experimentally assesses model assumptions (pPFR)*.

This work uses Eq (4) to simulate 1-minute ICP means from pulsatile ABP and CBF signals under constant CA processes, as in [23, 15]. This model is validated using shorter non-invasive CBF velocity measurements from the middle cerebral artery. Longer, non-pulsatile perfusion timeseries available to the present study require use via model inference (detailed in SI 1). The technique estimates CBF from CPP by optimizing non-negative control parameters *α*, ensuring consistency between Eq (4) and CPP data at each step.

#### 2.2.1 Posterior correction and hypothesis assessment

Model-estimated CBF is recalibrated to account for uncertainty of model-to-observation CBF correspondence and to test the asserted PFR hypothesis. Global CBF estimates (in mL/s) must be compared with local perfusion data (in mL/hg/min) by overcoming personalized biases and scale differences arising from patient-specific anatomy, injury, and probe location. Scale and bias are adjusted by fitting CBF estimates to perfusion data with a linear correction *m ·* CBF + *b*. Note that *m* is identified by minimizing the root mean squared error (RMSE) between CBF and perfusion scaled by their respective variances so that *m* is normalized. The *sign* of the correction slope *m* identifies whether the model-assumed pPFR is correctly asserted and is not affected by normalization.

Positive slopes (*m >* 0) signify CA impairment with perfusion data aligned with pressure-passive CBF assumptions (pPFR). Negative slopes (*m <* 0) signify opposition of perfusion data to these assumptions and more likely arising from nPFR-driven dynamics. Near-zero slopes identify no clear net CPP-CBF relationship over the 2-hour experiment. PFRs, or dynamical regimes, are categorized by the slope *m* of the correction for each experiment: pPFR cases are identified by *m >* 0.2; nPFR by *m <* 0.2; and zPFR by *|m| ≤* 0.2.

#### 2.2.2 Mean flow index calculation

Mean flow (Mx) index quantifies CA by gauging the relation of CBF changes to those CPP. The averaging and correlation windows affect the timescale and resolution of Mx, and there is no consensus choice for these parameters[33]. This work calculates Mx using 6-minute windows defined by 30-sample correlations of 12-second averages of CBF and CPP data. The choice stabilizes the index [29] while incorporating sub-minute pressure-flow variations inaccessible to the minute-scale model.

### 2.3 Experiment Selection

Experiments in this work aim to characterize PFRs by analyzing numerical simulations, clinical indexes (Mx and PRx), patient observations during intervals satisfying data requirements. From the available patient data, fourteen patients were identified with jointly recorded ABP, ICP, and perfusion; SI 2) summarizes the epochs containing the required data. Intervals of 100–140 minutes (nominally, 2 hrs) were algorithmically extracted from these patient records by excluding periods of missing data while ignoring gaps in perfusion data up to 10 minutes that may result from probe calibration. The extraction identified 193 patient-data intervals, which were further screened to omit those with perfusion sensor data flagged as faulty or where signals violated quality control thresholds. This filter removed intervals where: (i) ABP is negative or identified as an outlier among the 99.8th percentile of the data; (ii) perfusion is negative, exceeds 130 mL/hg/min globally, or has a 5-minute mean over 95 mL/hg/min; or (iii) ICP is negative anywhere or continuously exceeds 100 mm Hg for longer than 5 minutes. The set remaining defines 83 2-hour intervals from 11 patients; these are used for computational experiments (SI 3). The 2-hour duration maximizes the likelihood of a posture change or suctioning occurring within each experiment; such events occur in the data with a median frequency of about 1.5 hrs. These disturbance events prompt hemodynamic responses that facilitate PFR identification.

## 3 Results

This work extracts, checks, and contextualizes assumed ICHD relationships to gain understanding that may improve care of TBI patients. It identifies PFRs for individual patient-intervals, providing additional information about patient hemodynamics and autoregulation status. Section 3.1 examines hemodynamics data and clinical indices with an emphasis on pressure flow relationships over timescales of days. Section 3.2 categorize apparent PFRs of the data over 2-hour patient-interval experiments using model simulated CBF. Section 3.3 assesses relationships between dynamics, indices, and model estimation for the numerical experiment intervals. The closing results section briefly synthesizes results into main conclusions.

### 3.1 Data-oriented analysis of dynamics via indices

Tabulated statistics of PRx and Mx over day-scale data (SI Table SI 2) suggest differences in PFR regimes as measured by the CA functionality indices. The Mx mean values are negative in only 6 patient-epochs, while the median value is *∼*0.36. This median Mx exceeds the threshold of Mx=0.3 commonly identifying impaired CA [12, 33] and suggests a statistical dominance of a pressure-driven flow (pPFR) among these records. However, over one quarter of the patient-epochs show decoupled perfusion and CPP (*|*Mx*| <* 0.15) when summarized over tens of hours. This suggests that CA is functional in a sizable portion of cases, although variability is high. The PRx values associated with neutral Mx values (PRx = 0.08) are significantly smaller (one-sided *p* = 0.007) than the remainder (PRx = 0.28). These neutral values indicate active CA influences when most data are examined over timespans of hours or less.

### 3.2 Model-estimated CBF

Eighty-three experiments simulated CBF from ABP and ICP data using a model to enforce positive pressure-flow association (pPFR) under CA acting at 1-minute timescales. Validity of experiment hypotheses are identified from the orientation of a signed correction and the consequent error reduction during the full 2-hour period. Correction parameters were determined by calibrating experiment CBF estimates to observed perfusion. Among the 83 experiments representing 11 patients, the distribution of PFRs categories is as follows: 46% were identified with pPFR, 28% with zPFR, 27% with nPFR. SI 3 chronicles experiment results and the associated summary (SI 4) shows these proportions to be independent of probe quality diagnostics.

#### 3.2.1 Experiments identified with positive PFR

Overall positive coordination between CBF and CPP (pPFR) is dominant in 38 of 83 experiments (46%). Cases identified by pPFR cases have CBF changes that strongly track the changes in the pressure gradient (CPP), which is detected by a positive correction slope *m >* 0. Fig 2 depicts three such examples that exemplify pPFR: perfusion and CPP are positively correlated, as are perfusion and estimated CBF. Model estimated CBF (panels a and c, lower) correctly estimates the form of the perfusion trajectory from CPP. Trajectories may include transient excursions and variability, which are visible in the evolution of clinical indices (Mx, PRx) (right).

**Figure 2:**
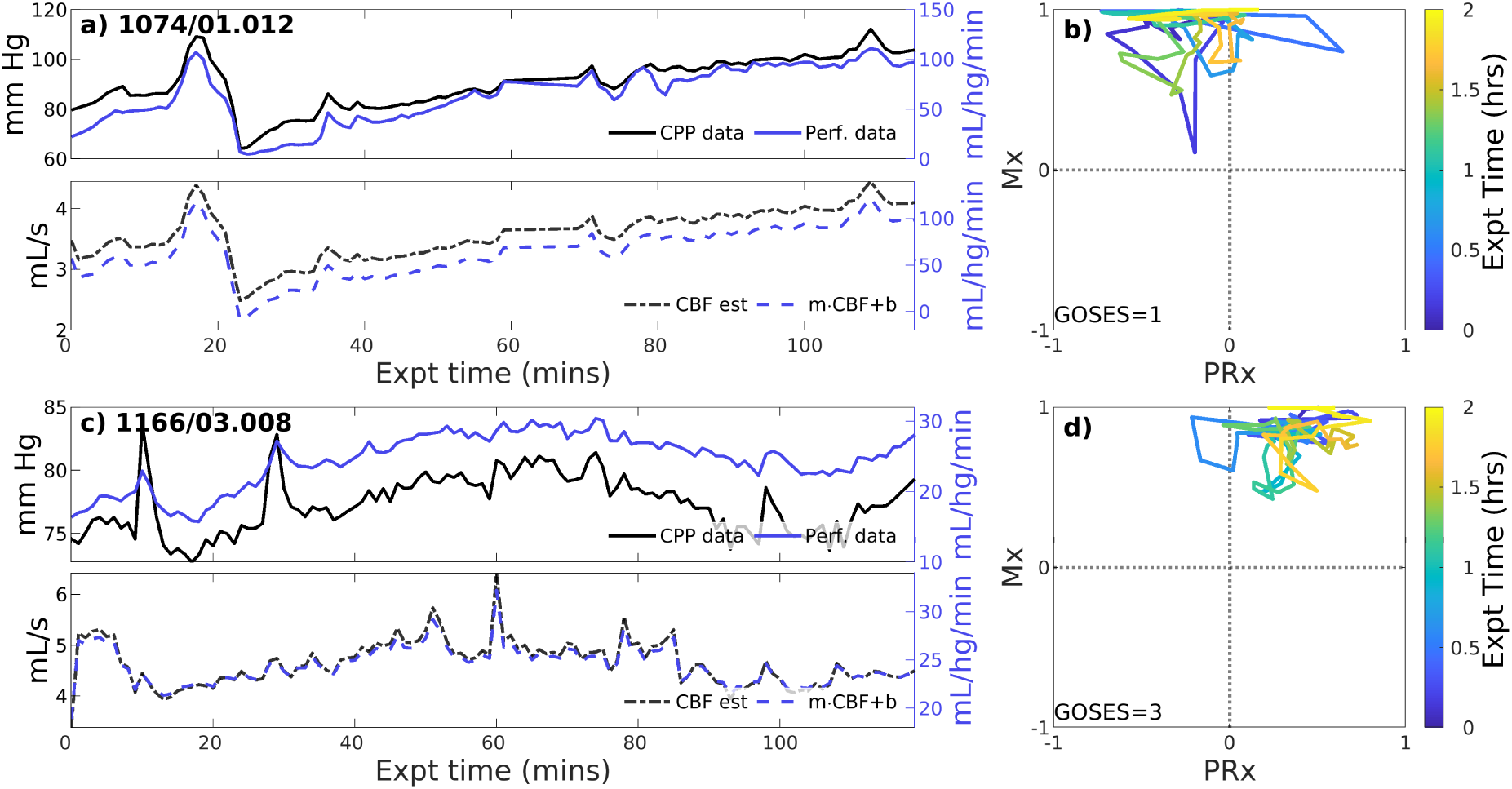
Two example experiments with positive PFR. (upper panels of a,c) Experiments in this set show consistent, positive coordination between CPP (black) and perfusion (blue) observational data. (lower panels) Model estimated CBF (black, dashed) follows changes in CPP; posterior correction (blue, dashed) preserves the original orientation of the estimate without a change in sign. (panels b,c) The associated trajectories in correlation indices show strongly positive Mx values, while PRx may be variable including sign changes.

#### 3.2.2 Experiments identified with zero-PFR

CBF-to-CPP coordination with an approximately zero-net mean is identified in 23 of 83 (27.7% of) experiments. These cases reflect statistical near-equilibrium between positive and negative PFRs, which may or may not result from decoupling of pressure and flow by active CA processes. Fig 4 illustrates several experiments where CPP (a and c, upper, black) has an irregularly structured relationship to perfusion data (blue) at both short and longer-term scales. CPP and perfusion data (upper panels) evolve independently at times and may include periods of both positive and negative coordination. CBF estimated from CPP cannot be corrected to consistently agree with perfusion data in these cases.

**Figure 3:**
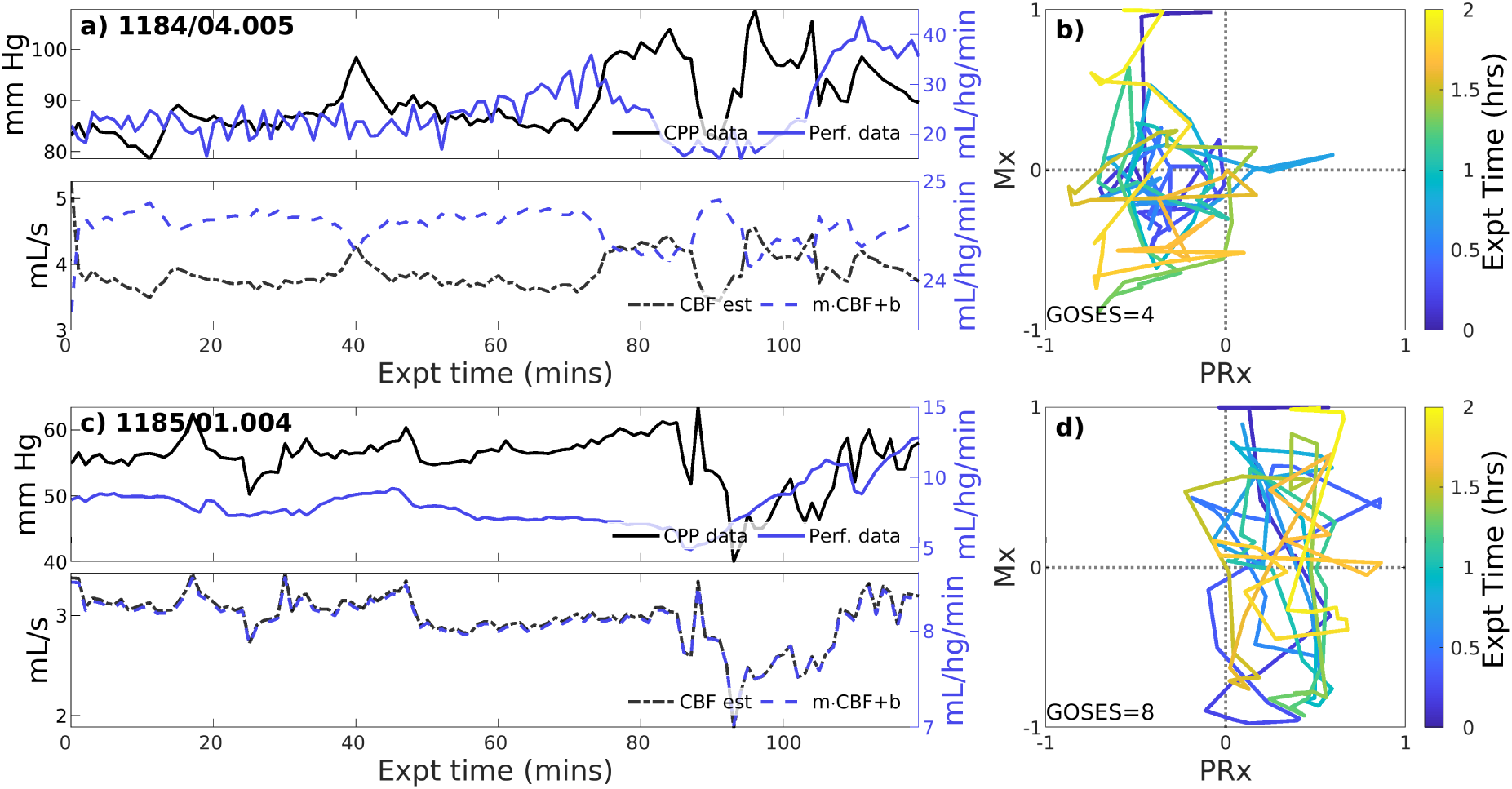
Two example experiments with zero PFR. The layout is the same as the previous figure. Pressure (solid black) and perfusion (solid blue) data do not consistently coordinate over the 2-hour experiment (upper panels of a,c). As a result, no suitable choice of Control parameters, corresponding to CA mechanisms, must alternate between positive and negative values to simulate these data for which no single PFR hypothesis suffices.

**Figure 4:**
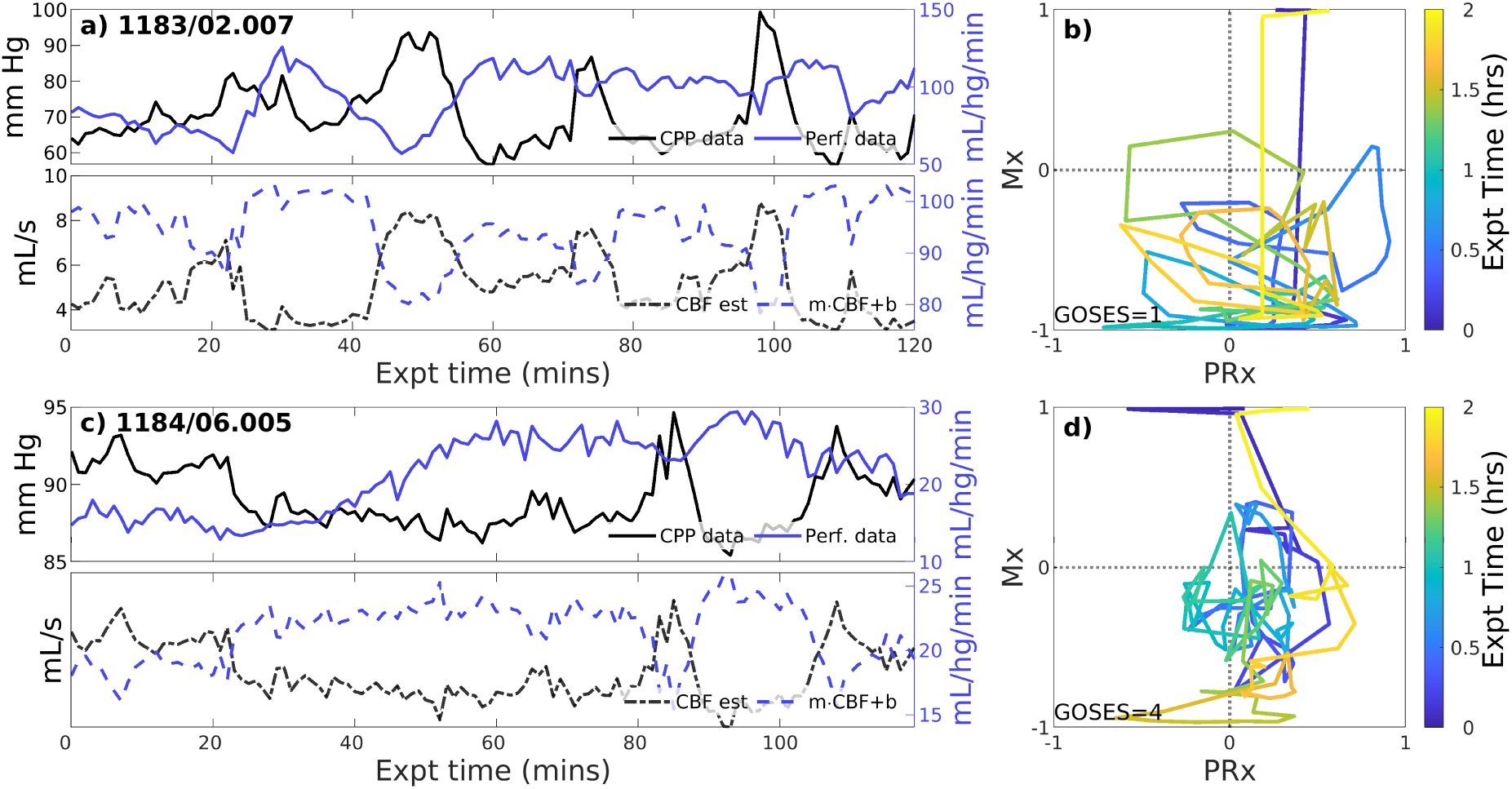
Example experiments with negatively-signed PFR. The layout is the same as the previous figure. Negative coordination between CPP (blue) and perfusion (magenta) observational data persists throughout many 2-hour experiments. Model estimated CBF (blue,dashed) follows changes in CPP and *requires* a sign change in the posterior correction (dashed blue) match observed perfusion. Both Mx and PRx can be highly variable in these cases whose underlying causes are not known.

#### 3.2.3 Experiments identified with negative PFR

Negative CBF-to-CPP coordination dominates 22 of 83 (26.5% of) experiments. Two examples (Fig 4) illustrate persistently negative correlation between observed CPP (solid black lines) and perfusion (solid blue) at both short and longer-term scales. Such behavior violates pPFR-oriented model hypotheses so that estimated CBF (dashed black) requires correction with a *negative* slope (‘*flipped* and scaled’) to correctly represent perfusion. Several investigated EHR records (SI 5) suggest reasons for nPFR appearance at hour timescales include metabolism and sedation, blood volumetric influences, and transitions across the limits of CA functionality.

### 3.3 ICHD data in relation to experiments

Analyzing observational data through the lens of model-identified PFRs can better characterize ICHD and associated properties. Fig 5 empirically establishes the connection between PFRs and CA metrics, Mx and PRx. The aggregated joint (PRx, Mx) distributions for pPFR and non-pPFR experiments (left and center) differ significantly (two-sample K-S test [50], *p <* 0.001, *D ≈* 0.335). This difference (right) suggests a familiar, interpretable discrimination of PFRs using CA indexes. Particularly, pPFR experiments are delineated by joint (Mx, PRx) values (right, blue line), generalizing the independent thresholds on Mx and PRx for CA function (green lines). Note that parameters defining the linear discrimination depend on the window scheme used in Mx and PRx calculation.

**Figure 5:**
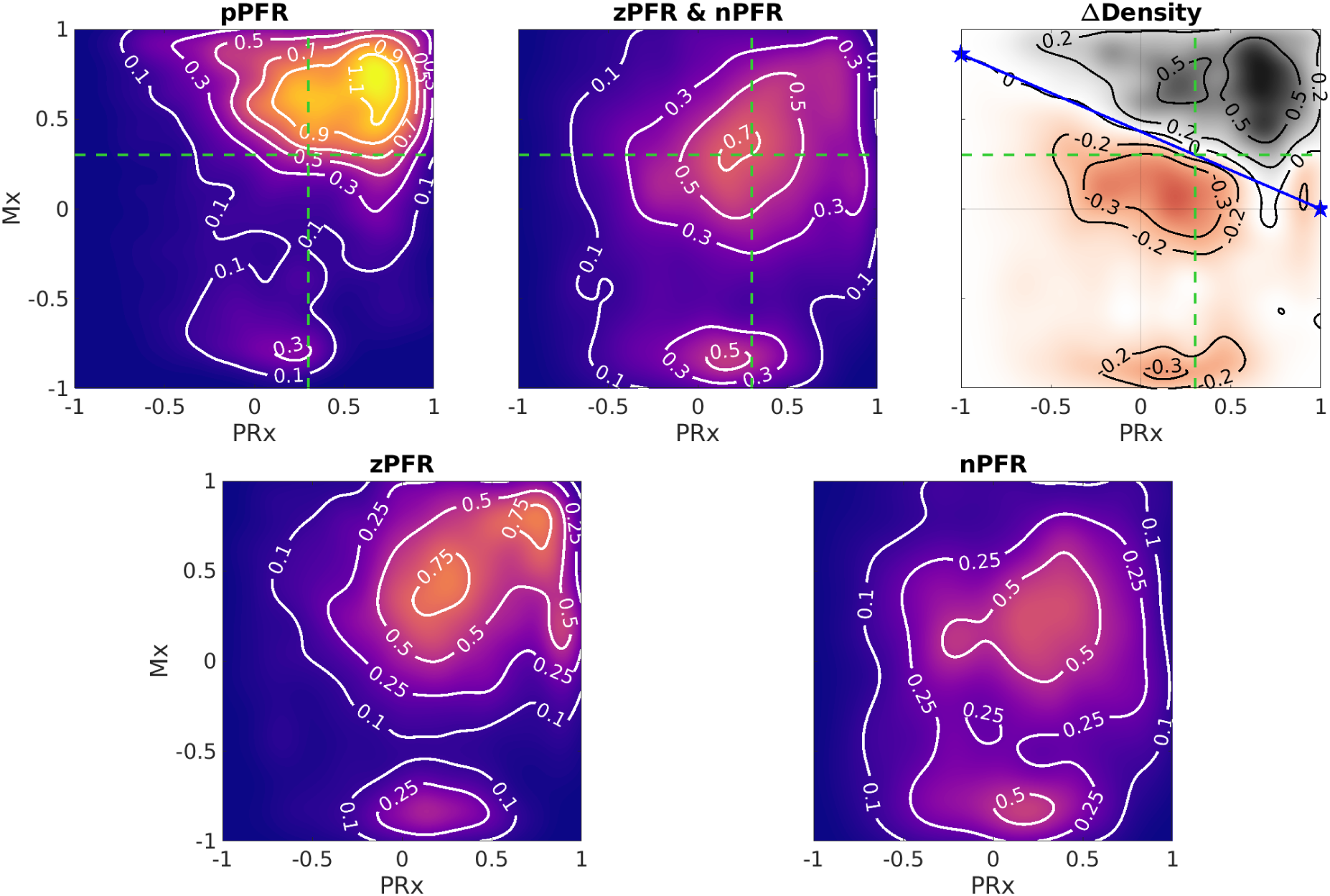
Differences in joint Mx and PRx distributions. Contours show distribution of (PRx, Mx) for pPFR (27.3K points), zPFR (13.6K points), and nPFR (15.8K points) experiments, excluding unqualified data (PPA*≥*5). For pPFR, the data density is highest at (PRx, Mx) = (0.7, 0.75) where both pressure and flow aspects of autoregulation are impaired. In the top right panel, pPFR experiments dominate the region Mx = 0.43 *·* (1 *−* PRx) comprising high positive values of PRx and Mx. Green lines represent current index thresholds of 0.3 above which CA is assumed to be impaired; delineation based on PFR is a consistent joint consideration of those thresholds. The corresponding zPFR and nPFR distributions are distinct (bottom row). The zPFR data include both strongly positive and negative Mx values, whereas nPFR data consist of near-neutral positive Mx and strongly negative Mx.

High Mx values associated with pPFR correspond precisely to hypothesis of impaired CA, while remaining experiments explain the central mass of data around (PRx, Mx)=(0.3, 0.3). The data associated with zPFR (bottom left) and nPFR (bottom right) experiments are also distinct, with the former being more variable and including both high and low values of Mx. This supports the hypothesis that longer-time decoupling of signals results from a balance of local positive and negative pressure-flow coordination which may be extreme in either direction. However, the nPFR subset (bottom right) lacks the presence of strongly positive Mx, so that their distribution includes a broad, neutral center near (PRx, Mx)=(0.27, 0.2) and a more extreme negative center near (0.152*, −*0.82). Within nPFR-identified data, the relative proportion occupied by the Mx-negative center decreases greatly when using Mx longer averaging windows, which suggests the involvement of processes occurring at sub-minute scales.

Identifying PFRs as above depends on CBF/perfusion data and it is desirable to seek characterization PFRs in more common data. For example, short-timescale autoregulatory responses to extra-hemodynamic influences [5] might be detectable when CA is functional within non-pPFR cases. However, variability of dynamics limits discriminatory data analysis because joint distributions of measurements are broad over 2-hour intervals. Characteristics of PFRs are instead pursued among observed states they associate with. Perfusion is excluded from the following analysis due to its rarity and dependence of its values on patient– and placement-specific properties.

PFR classification over 2-hour windows summarizes significant short-term variability at 1-minute timescales. Figure 6 shows the corresponding distributions of ICHD (Figure 6, a–d) and non-hemodynamic variables (e– h). All distributions except ABP are distinct (pairwise two-sample KS tests, *p <* 0.005) and show significant differences in medians (pairwise two-sample t-tests, *p <* 0.05) except: nPFR and pPFR have similar ICT, and SpO_2_ is tightly centered near 100% across all three groups. Positive PFR data indicate elevated ICP, reduced CPP, instances of reduced tissue oxygenation, and a majority of PRx values indicating impaired CA. The zPFR data link to moderate ICP and diminished CPP with moderate positive PRx centered near 0.3, the current threshold of CA function. The zPFR also have the highest median tissue oxygenation under similar CPP to pPFR cases, along with elevated ICT.

**Figure 6:**
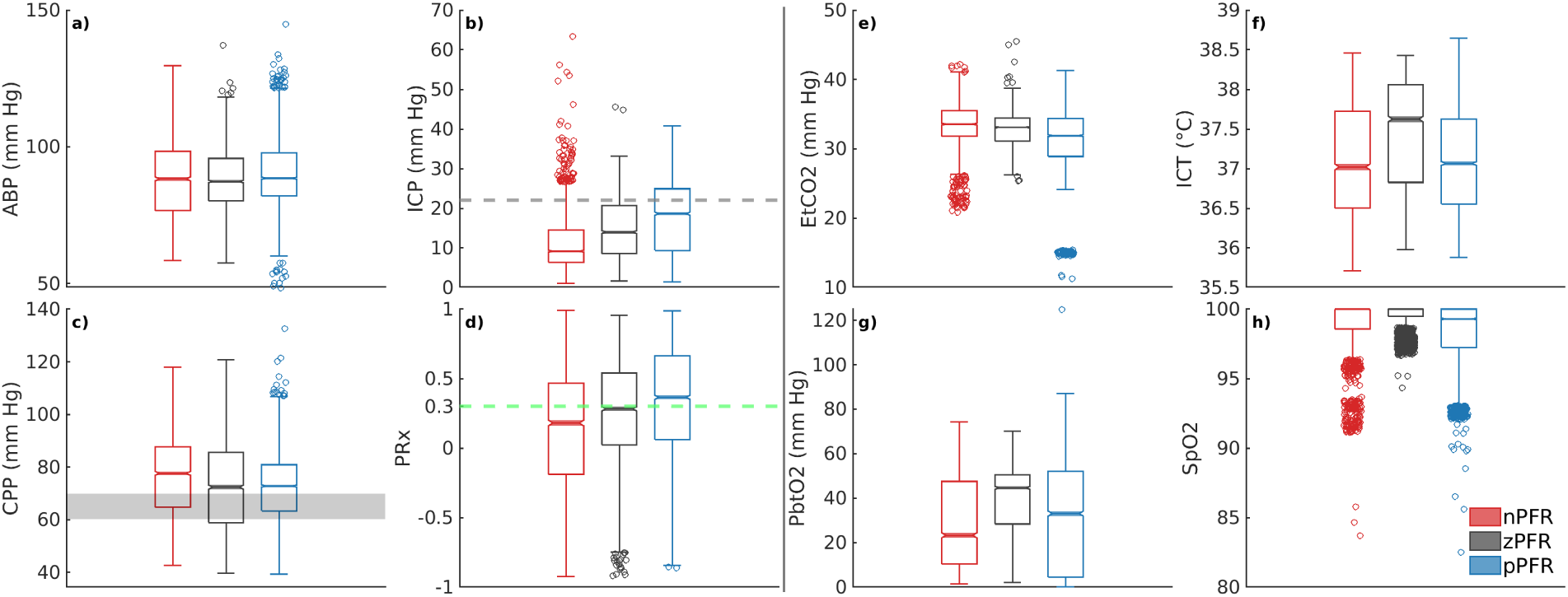
Distribution of 1-minute averaged data in PFR-identified experiments. In the left half, panels (a–d) show ABP, ICP, CPP, and PRx, respectively. In the right half, panels (e–h) show EtCO_2_, ICT, P_bt_O_2_, and SpO_2_, respectively. (c) CPP medians exceed the protocol target range of 60–70 mm Hg. Both brain tissue oxygenation (g) and ICT (f) are elevated in zPFR relative to other data, although this may also reflect differences in patient care during intervals selected for experiment. Fully characterizing nPFR likely requires finer temporal and patient-specific analysis to identify the respective influences of high and low ICP, EtCO_2_, both systemic and brain-tissue metabolic factors.

The nPFR data, in contrast, have lower PRx values that indicate functional CA and concentrate at both low and super-critical ICP: over 15% of nPFR ICP is hypertensive (*>* 20 mm Hg) while the median is *∼* 9 mm Hg. This multimodal structure suggest that nPFR comprises multiple sub-dynamics involving flow-related effects not quantified by PRx. Compared with outer cases, nPFR has reduced median tissue oxygenation paired with elevated CPP and the highest incidence of SpO_2_*<*92%. The data suggest nPFR characterization includes CA-mitigated flow to limit hyperemia (at low ICP/high CPP) and hypoxia (with low P_bt_O_2_ in response to further reduced SpO_2_). However, the 1-minute analysis considered here does not exclude volume-driven intracranial hypertension as a possibility in high ICP cases, as this requires finer temporal resolution to pinpoint causal influence.

### 3.4 Results Summary and Conclusions

Hypothesis-oriented experiments and PFR-differentiated data analysis found that:

1. Dynamics involving pressure-driven flow, pPFR, appeared throughout 46% of the experiment intervals (41 of 83) that were consistent with model assumptions. Remaining experiments divide about equally between nPFR (27%) and zPFR (28%) categories and include dynamics in violation of basic fluid mechanical rules governing pressure-driven flow. Hour-scale dynamics of zPFR cases typically feature an equilibrium between pPFR and nPFR modes with both high and low Mx values, rather than comprising locally decoupled signals.
2. Computational ICHD models fitting the pPFR domain extended to the nPFR domain with minor modification because their physical assumptions are mathematically equivalent up to a sign change. A similar approach may be pursued for zPFR cases composed from mixed pPFR/nPFR dynamics by changing assumptions (and model signs) coincident with changes in PFR.
3. Pressure-flow identities could be inferred from the data through clinical indices Mx and PRx. Most nPFR experiment data has PRx values below 0.15, while the majorities for pPFR and zPFR experiments were above 0.25. Highly variable dynamics of both zPFR and nPFR cases are captured in trajectories of Mx and PRx. PFR domains are delineated by joint consideration of Mx and PRx in a way that generalizes the current, independent thresholds for identifying CA impairment (Fig5, right).
4. Negative pressure-flow relationships (nPFR) can persist for hours and exist over multiple timescales including the full duration of many 2-hour experiments. Data in nPFR cases associate with both intracranial hypertension and low ICP; about 15% of nPFR data features ICP*>*20 mm Hg while half falls under 10 mm Hg under similar CPP. This is evidence of multiple nPFR sub-dynamics that plausibly discern between metabolically-driven flow at low ICP and volume-driven behavior at elevated ICP.
5. Detailed examination of patient trajectories (SI 5) suggests that state-dependent engagement of CA in addition to the metabolic environment (Fig6) affect the dominant PFR, and possible volumetric influences could not be ruled out. Sedation, mannitol, and patient stimulation events strongly influence the ICHD state and remain confounding factors in simulation-based experiments.

## 4 Discussion

This work analyzed a novel neurocritical dataset containing cerebral blood flow (CBF) and cerebral perfusion pressure (CPP) to investigate hour-scale pressure-flow relationships (PFRs). These relationships are essential, underlying components of computational and conceptual models of hemodynamics, and required rare, previously unavailable continuous perfusion or blood flow data to validate over longer scales. The positive pressure-flow relationship (pPFR), typically assumed in hemodynamics models and the mean flow index (Mx), was identified as dominant in 46% of the 83 2-hour experiments explored across 11 patients. About 28% of experiments showed no or weak orientation in PFR (zPFR) as expected when cerebral autoregulation (CA) is functional and engaged; such dynamics were not expected to be deterministically simulated. However, 27% of experiments were best described by a negative PFR (nPFR) not fitting physiologically justified dynamics. This behavior is of interest for pressure-guided TBI management because the common pPFR assertion fails in these cases.

The pressure-driven flow regime pPFR (Eq (1)) is tied to dynamics with dysfunctional, impaired, or un-engaged CA mechanisms because blood flow is driven primarily by the arteriovenous pressure gradient, CPP. The dynamics of zPFR (Eq (2)) include active decoupling of pressure and flow by cerebral autoregulation, so that neutral values of PRx (ABP to ICP correlation) and Mx (CPP to flow correlation) were anticipated. The nPFR cases featured neutral PRx and strongly negative Mx, while associated observational data (Fig6) suggests multiple possible origins including flow driven by metabolic demand and volume-driven pressure dynamics. The associated differences in joint (PRx, Mx) distribution (Fig5) also show that the PFR perspective aligns with and generalizes current thresholds for CA impairment. These developments aim to support improvement of mechanistic process models and refine clinical guidance from CBF observation.

### 4.1 Key findings

Assumptions of pressure-driven flow do not apply to negative PFR cases, which are characterized by *strongly anti-correlated CPP-CBF dynamics*. These dynamics are governed by distinct physiological processes and violate model assumptions in a precise, identifiable, and correctable way. Specifically, they reverse the orientation of predicted CBF amplitudes about the mean (see Fig 4), an error correctable by a strategic sign change in Eq (4). Specifying the correct hypothesis for these cases demands a more comprehensive understanding of the nPFR dynamics. The three PFR regimes were associated with considerable differences among peripheral data (*viz.* P_bt_O_2_, ICT, and SpO_2_) and parameters (PRx, Mx), although distributions were broad at both intra– and cross-patient levels. *Investigation of available data with numerous sources of heterogeneity could not establish PFR classification criterion without direct knowledge of CBF*.

The linear segmentation of (PRx, Mx) parameter distributions provided PFR-discerning criterion Mx *>* 0.43 *·* (1 *−* PRx) that qualifies CA impairment under the hypotheses of pPFR. This delineation (Fig 5) separates dynamical regimes in a way that generalizes current thresholds (of 0.3) for CA impaired based on Mx and PRx, highlighting the benefit of combined over individual index use. Such thresholds are sensitive to index calculation window and should be considered in relation to timescales of interest. The analysis also illustrates that PRx and Mx convey different information about ICHD: PRx better discriminates pPFR from other categories, while Mx best discriminates the nPFR data.

The pPFR-identified cases align with high PRx and Mx values, while cases associated with nPFR appear to contain multiple dynamics. Most zPFR data are associated with neutral PRx and Mx as CA and/or therapeutic interventions statistically decouple ABP, ICP, and CBF and extra-hemodynamic signals. Rule-based modeling of uncorrelated data is difficult, but the indication of CA function indicates little advantage to doing so for decision support. However, the anticorrelated CBF-CPP dynamics found in nPFR-labeled data are simulatable with a simple change in the model and hypothesis.

PFR-level differences could not be strongly established among signals independently of CBF, which suggests that proxy representation of CBF requires deeper consideration. The detection of possible metabolic influences on CBF within nPFR motivates the use of empirical modeling to assess *which* clinically observable variables best relate to quantities of interest. Fig 7 ranks the influence of MMM data as predictors of ICP and CBF predictors in 83 experiment intervals as determined by Gaussian process regression [52]. For empirically predicting ICP and CBF, the most influential signals relate to metabolism/infection, respiration, and sedation rather than hemodynamics (outlined in red). Both nPFR and pPFR qualitatively share this ordering, which suggests their variables of interest might be approximated by common model framework incorporating extra-hemodynamic factors over 1–2 hour timescales.

**Figure 7:**
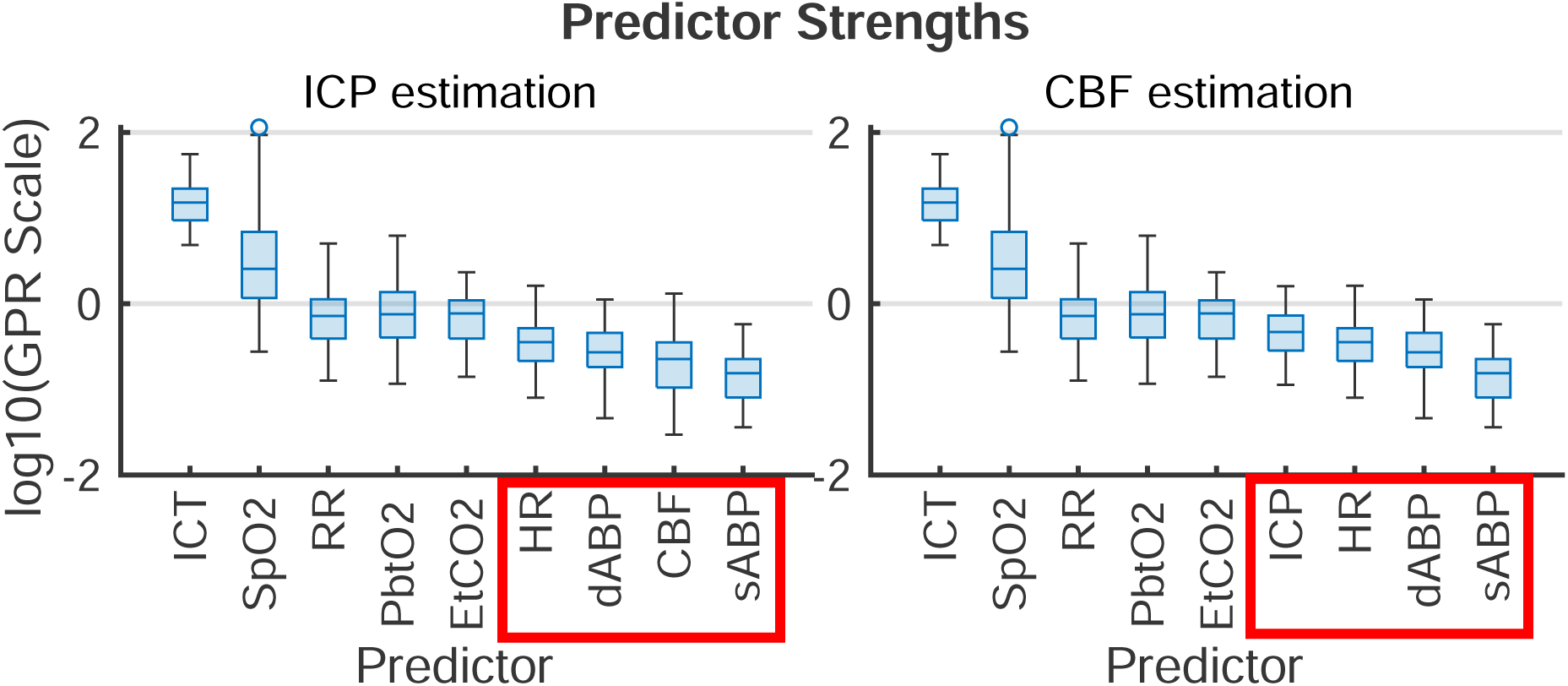
Gaussian process regression fitting of ICP (left) and CBF (right) from 1-second averaged data across the 83 2-hour experiment intervals. Box plots give the distribution of scales of predictor influence on a logarithmic scale ordered decreasingly by median. ICHD model variables, indicated in red boxes, have low empirical predictive rank over the 2-hour timescale. Importance in the fitting is found by automatic relevance determination (ARD, [28, 30]) using kernel function parameters associated with predictor lengthscales. The ranking is robust under alternate regression strategies (*e.g.*, Lasso and Ridge Regression).

**Figure 8:**
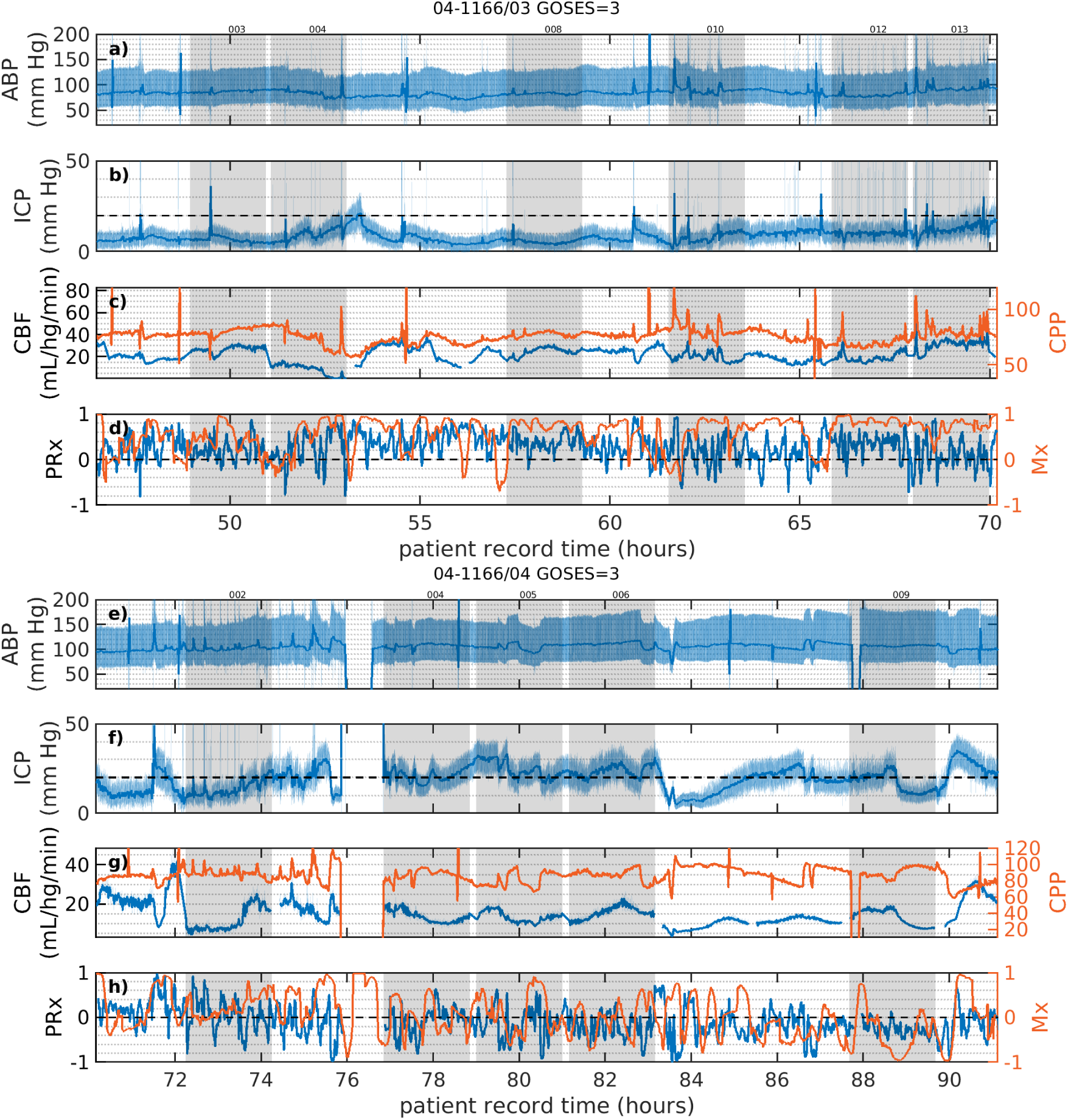
Two epochs of longitudinal data for patient 04-1166 contextualize PFR evolution and numerical experiments. Panels a–d and e–h present timeseries of from epochs 3 and 4 corresponding to summary table rows 24 and 25, respectively. Left axes and blue curves correspond to MMM data of patient 1166 during the *∼*2-day period starting 46.5 hours into hospitalization. Dashed line in the second rows indicates 20 mm Hg ICP. Grey boxes throughout denote experiments: the last four experiments in the upper panel (rows43–46 of experiment tables) are pPFR-governed while all experiments in the lower panel indicate nPFR. Experiment 008 in panels a–d corresponds to the pPFR example of Fig2c,d.

### 4.2 Limitations and Implications

Numerical simulations in this hypothesis-based analysis rely on specific data and data quality, potentially biasing the representation of patients with more abundant, complete, and clean data who may not reflect PFRs in the entire patient population. However, high intra-patient variability and long data series led to multiple PFRs appearing in most patients without a discernable pattern over time. A robust analysis of temporal patterns in PFR warrants further study; the 166 patient-hours categorized from the 67 patient-days of data are insufficient to address this topic here. The lack of population generalizability does not impair the study objective of validating model assumptions, rather than PFR distributions or a specific model implementation.

The presented PFR identification depended on ad hoc linear correction to equate global CBF estimates with local tissue perfusion data. This adjustment addressed measurement uncertainties and patient data variability while preserving signed coordination between pressure and flow. However, the true CBF-perfusion relationship is more complex and likely nonlinear as it depends on interacting factors including vascular stiffness, Circle of Willis geometry, and blood flow distribution at the probe site. A dedicated study com-paring localized thermodiffusively-measured perfusion to global CBF estimates (via transcranial Doppler measurement) at the middle cerebral artery remains important, particularly to rectify the influence of injury distribution and probe location on observed flow. Although perfusion data were screened for quality based on probe diagnostics, unidentified factors such as spurious trends caused by ABP, ICP, or CBF sensor drift cannot be completely dismissed. Minimal impact on the primary findings of this study is expected as it is improbable that such artifacts occurred consistently over hour-long timescales of multiple patients.

The pPFR, defined by known physiological mechanics, served as a natural null hypothesis for PFR classification along with zPFR, whose hemodynamics include pressure-independent CBF. However, half of zPFR-identified experiments showed highly variable alternation between nPFR and pPFR extremes (Fig3c– d) rather than the more stable balance (a–b). Subintervals of some highly variable zPFR cases may be categorized as nPFR or pPFR, making them potentially modelable (discussed below) but partially unexplained. Analysis identified that nPFR, the alternative hypothesis to positive and negative coordination, comprises dynamics that diametrically opposite the pPFR assumptions used in models and underlie Mx as a clinical measure of CA function. Chart review (SI 5) of several nPFR cases suggests related factors include crossing limits of CA, hyperemia, and the effects of intervention and stimulation. Despite an unclear characterization of these nPFR dynamics through modeling and analysis of data, this work identifies an extension of existing assumptions to model and reason about them.

### 4.3 Modeling Implications

ABP-only estimation of ICP [44] generated consistently erroneous trends and phase of ICP in some experiments. Error diagnosis could not be explained without perfusion data and motivated this work. Although the errors were correctable by negating the pressure-flow relationship (a sign change in Eq(4)), such modification contradicted physical assumptions of the model. Change of model hypothesis are now justified by examining MMM data of experiments: a sign change in Eq(4) is necessary for dynamics governed by nPFR rather than pPFR. The change specifically addresses nPFR cases where dynamics are opposite to the pressure-driven flow assumed by the model. Physiologically, the correction corresponds to CA modulating flow opposite to CPP changes; its computational counterpart in the compartmental model framework are negative resistances. This amended hypothesis pragmatically extends the pressure-driven flow model framework to nPFR dynamics, despite being incorrect for *e.g.*, volume-driven pressure or metabolically-driven flow regimes.

Harmonizing nPFR and pPFR assumptions into a common framework requires a method for correctly choosing which hypothesis applies for a particular patient, state, and time. However, a mechanistic representation is complicated by the distinct nPFR sub-dynamics and extra-hemodynamic CA stimuli that remain incompletely characterized. An accessible alternative is to wrap the current implementation in a machine learning (ML) layer [16, 27] which predicts the PFR and necessary sign changes from training data and other relevant streams of patient observation. This approach leverages existing mechanistic ICHD models and their modification to extend model applicability to clinically-relevant, real-world cases.

### 4.4 Practical Implications

CPP/ICP-oriented therapies rely on assumptions valid under pPFR that may not be valid for nPFR patient dynamics. Within zPFR dynamics where functional CA modulates flow, manipulation of CPP is only likely to directly influence CBF if CA response is altered, by *e.g.* by moving pressures across the limits of CA function. In nPFR dynamics, however, use of CPP to proxy CBF [14, 1] incorrectly assumes positive pressure-flow coordination. ICP/CPP-based therapies may not ensure adequate perfusion in nPFR dynamic regimes which include flow/volume-driven pressure and metabolically-driven flow. In the former, interventions that manage intracranial volume budget (CSF drainage, heartrate) could be effective strategies to optimizing perfusion rather than raising ABP, particularly if the blood-brain barrier is compromised [31]. In the latter, which are postulated to feature low ICP (Fig6b), CPP manipulation may unnecessarily disrupt endogenous flow regulation.

The ICHD system variables (CPP, CBF and perfusion, and the set of CA functions) warrant careful treatment of causal relationships, which can be overlooked when analyzing states rather than trajectories. Mx and PRx are CPP-CBF and ABP-ICP correlations, respectively, which measure different functional aspects of CA without causal direction between variables. Considering patient trajectory through the PFR perspective offers a more granular, dynamical perspective and incorporates causal logic into ICHD relations in contrast to separate use of Mx and PRx. Unlike pressures that influence the intracranial volume budget, blood flow contributes to it directly. Observations of CBF changes in the context of the pressure environment may also improve metrics and targets for TBI management that account for volume (*viz.* the Lund concept [17] and implicitly CPP_opt_ [13]). More long-time observation of CBF is required to investigate whether CPP/ICP-oriented manipulations result in appropriate perfusion under nPFR dynamics when CPP fails to proxy blood flow. The PFR perspective may also be valuable for discriminating which patient ICHD behaviors benefit from vascular protection by ICP-targeting therapies vs. flow-optimizing CPP-optimizing targets, as [20] found management outcomes depended on patient autoregulatory state.

### 4.5 Concluding Remarks and Future Work

The baseline assumption of positive pressure-flow coordination affects neurocritical care of TBI patients because ICP– and CPP-targeting guidelines aim to ensure adequate brain tissue perfusion through pressure management. This work analyzed those underlying assumptions for consistency with against continuous, invasively-monitored TBI patient data, employing a physiological model to classify pressure-flow relationships based on assumption appropriateness. It revealed ICHD categories explained by whether CA processes were functional and found additional dynamics with strongly negative pressure-flow coordination sustained over long timescales. A simple yet unanticipated modification of assumptions, corresponding to negative pressure-flow coordination, was required in these latter cases. The necessity explains the inaccuracy of CBF-less ICP model predictions that motivated this study, but it implies a similar misconception may exist in TBI management through pressure targets alone. This conclusion motivates use of CBF observation for personalizing neurocritical care to CBF and other observations. As PFRs distinguish hemodynamics by incorporating pressure and flow, and therefore generalize Mx and PRx (Fig 5), they are means toward that development. The mathematical formulae in ICHD models can be trivially modified to account for nPFR dynamics without understanding the mechanisms underlying their origin. Future work identifying the causes and physiological mechanisms of nPFR will further refine PFR characterization and improve reliability of predictive decision support tools for cases currently outside the model framework.

## Declarations

### Ethics

Not Applicable

### Consent for publication

Not Applicable

### Authors’ Contributions

Co-author contributions to this research are as follows: JS – conceptualization, development, design and analysis of experiments, interpretation of results, and drafting; BF – funding, collection, quality control, and migration of data, interpretation of results, and providing critical feedback; TB – funding, critical guidance of conceptualization and drafting, and manuscript feedback; JB – conceptualization, interpretation, and critical feedback on experiments and the manuscript; SP – providing critical feedback and editing the manuscript; and DA – funding, conceptualization, interpretation, manuscript feedback, and general project oversight.

## Acknowledgments

Authors thank Meg Rebull supporting efforts and to Jan Claassen of Columbia University for delightful discussions about the final sections.

## Data Availability

Data used in this work comprise high frequency timeseries and waveforms gathered during prospective clinical trial (https://tracktbinet.ucsf.edu/). Data access may be formally requested through the TRACK-TBI program at https://tracktbi.ucsf.edu/collaboration-opportunities.

## A SI 1

Assessing the internal consistency and effectiveness of CBF estimation requires numerical experimentation. However, the mechanistic relationship of CBF to the applied pressure gradient (*∇p*, the continuous-time equivalent of CPP) involves the unknown effects of CA. Simultaneous estimation CA and CBF is possible via inverse modeling centered on observed ABP, (*P* ^obs^). Optimization sequentially identifies CA parameters (*α*) within 1-minute windows until the model ICP estimate (*I*) agrees with observed ICP (*I*^obs^) as measured by mean squared error (Eqns(5) below). The calculated CBF estimate (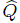) is the optimal CBF (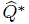) in the model framework in that it reflects flow under a well-approximated pressure gradient via strategically optimized CA parameters (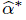). Although implemented in discrete time, the continuous-time optimization can be expressed as 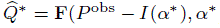) for **F** of Eqn(4) where

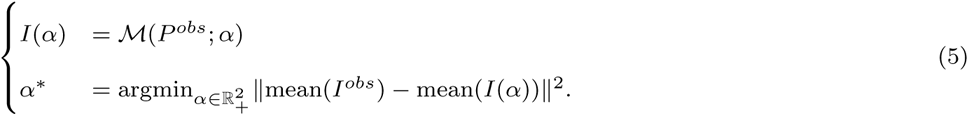

Experiment definition here requires continuous input of ABP data for forward-time propagation, ICP data for model inversion, and CBF data for posterior correction and error diagnosis.

## B SI 2

Folder level summary of ICHD content of data in University of Cincinnati cohort of patients with recorded invasive regional perfusion (ml/hg/min), ABP (mm Hg), and ICP (mm Hg). Time *t* s recorded in hours. PRx is calculated bedside on Moberg CNS monitor platforms using 30 samples of 10-second averages of ICP and CPP. Mx is calculated as part of this analysis using 30 samples of 12-second averages of perfusion and CPP. One epoch with less than 1 hour of valid perfusion data is omitted.

| $k$ | Data | GOSE | $t$ | ABP | ICP | Perf | PRx | Mx |
| --- | --- | --- | --- | --- | --- | --- | --- | --- |
| 1 | 1073/01 | 1 | 0.0–9.5 | 80.2 ± 15.1 | 27.0 ± 6.0 | 11.9 ± 5.8 | 0.49 ± 0.4 | 0.62 ± 0.4 |
| 2 | 1073/02 | 1 | 9.8–18.5 | 85.9 ± 10.8 | 28.5 ± 5.8 | 46.0 ± 25.3 | 0.47 ± 0.4 | 0.40 ± 0.6 |
| 3 | 1073/03 | 1 | 18.6–34.0 | 86.6 ± 11.7 | 22.4 ± 7.2 | 85.3 ± 11.4 | 0.43 ± 0.4 | 0.01 ± 0.4 |
| 4 | 1073/04 | 1 | 34.1–38.0 | 97.1 ± 13.5 | 15.9 ± 8.4 | 149.2 ± 4.0 | –0.16 ± 0.3 | 0.40 ± 0.1 |
| 5 | 1074/01 | 1 | 0.0–22.1 | 98.3 ± 26.0 | 19.0 ± 3.9 | 33.8 ± 41.5 | 0.12 ± 0.4 | 0.43 ± 0.4 |
| 6 | 1074/02 | 1 | 22.2–45.5 | 106.2 ± 11.9 | 23.9 ± 3.5 | 33.4 ± 22.0 | 0.12 ± 0.3 | 0.58 ± 0.3 |
| 7 | 1075/02 | 1 | 8.8–18.3 | 76.1 ± 10.1 | 17.5 ± 3.8 | 96.8 ± 25.0 | –0.03 ± 0.3 | 0.03 ± 0.2 |
| 8 | 1075/05 | 1 | 57.1–99.0 | 93.3 ± 13.6 | 15.8 ± 4.8 | 1.7 ± 5.2 | –0.04 ± 0.3 | –0.02 ± 0.3 |
| 9 | 1086/01 | 1 | 0.0–25.3 | 83.3 ± 10.0 | 26.3 ± 6.2 | 29.9 ± 12.5 | 0.31 ± 0.3 | 0.64 ± 0.3 |
| 10 | 1086/02 | 1 | 25.4–28.1 | 91.3 ± 12.7 | 40.8 ± 6.2 | 40.4 ± 7.8 | 0.48 ± 0.4 | 0.45 ± 0.4 |
| 11 | 1086/03 | 1 | 28.4–32.4 | 96.7 ± 10.4 | 35.3 ± 8.8 | 45.8 ± 15.2 | 0.40 ± 0.3 | 0.71 ± 0.3 |
| 12 | 1086/04 | 1 | 32.5–47.4 | 93.3 ± 9.7 | 26.6 ± 8.9 | 33.9 ± 20.5 | 0.14 ± 0.3 | 0.75 ± 0.3 |
| 13 | 1086/05 | 1 | 47.4–71.2 | 92.4 ± 10.5 | 26.8 ± 6.3 | 37.7 ± 14.5 | 0.39 ± 0.4 | 0.55 ± 0.4 |
| 14 | 1086/06 | 1 | 71.3–96.2 | 88.4 ± 11.4 | 28.5 ± 4.2 | 30.9 ± 13.1 | 0.60 ± 0.3 | 0.58 ± 0.3 |
| 15 | 1086/07 | 1 | 96.4–100.0 | 104.7 ± 10.6 | 35.8 ± 3.3 | 35.5 ± 16.3 | 0.46 ± 0.3 | 0.53 ± 0.3 |
| 16 | 1086/08 | 1 | 100.7–107.7 | 62.8 ± 52.1 | 38.3 ± 5.0 | 17.8 ± 6.7 | 0.42 ± 0.4 | 0.09 ± 0.5 |
| 17 | 1140/01 | 5 | 0.0–94.8 | 79.1 ± 14.6 | 13.7 ± 4.9 | 14.5 ± 10.4 | 0.02 ± 0.4 | 0.25 ± 0.4 |
| 18 | 1141/01 | 1 | 0.0–105.4 | 79.1 ± 14.8 | 18.0 ± 4.5 | 13.0 ± 9.4 | 0.46 ± 0.4 | 0.29 ± 0.4 |
| 19 | 1160/01 | 1 | 0.0–24.1 | 83.5 ± 17.0 | 11.3 ± 2.9 | 22.0 ± 12.4 | 0.38 ± 0.3 | 0.45 ± 0.3 |
| 20 | 1165/01 | 1 | 0.0–17.1 | 88.8 ± 14.8 | 23.1 ± 6.2 | 22.0 ± 6.6 | 0.48 ± 0.3 | 0.36 ± 0.4 |
| 21 | 1166/01 | 3 | 0.0–25.7 | 84.4 ± 12.8 | 6.7 ± 4.2 | 25.7 ± 10.9 | 0.22 ± 0.3 | 0.46 ± 0.3 |
| 22 | 1166/02 | 3 | 25.7–46.5 | 94.0 ± 11.2 | 8.4 ± 4.7 | 23.4 ± 8.1 | 0.34 ± 0.3 | 0.46 ± 0.3 |
| 23 | 1166/03 | 3 | 46.5–70.2 | 84.6 ± 9.2 | 8.7 ± 4.4 | 21.7 ± 8.0 | 0.27 ± 0.3 | 0.54 ± 0.3 |
| 24 | 1166/04 | 3 | 70.2–91.3 | 104.4 ± 14.5 | 19.1 ± 7.2 | 14.8 ± 6.1 | –0.12 ± 0.3 | 0.18 ± 0.4 |
| 25 | 1171/02 | 1 | 0.7–86.1 | 82.2 ± 13.6 | 8.2 ± 5.9 | 42.9 ± 2.9 | 0.13 ± 0.5 | 0.95 ± 0.2 |
| 26 | 1178/01 | 3 | 0.0–52.8 | 84.6 ± 12.1 | 14.1 ± 3.4 | 47.0 ± 14.5 | 0.14 ± 0.3 | 0.12 ± 0.5 |
| 27 | 1183/02 | 1 | 25.2–94.3 | 86.2 ± 14.7 | 13.5 ± 7.4 | 80.8 ± 36.2 | 0.08 ± 0.4 | –0.58 ± 0.4 |
| 28 | 1183/03 | 1 | 106.3–133.5 | 96.4 ± 11.5 | 15.2 ± 8.8 | 62.8 ± 25.6 | –0.00 ± 0.3 | –0.36 ± 0.4 |
| 29 | 1183/04 | 1 | 133.5–136.9 | 78.9 ± 40.9 | 14.8 ± 6.0 | 69.6 ± 12.1 | –0.18 ± 0.3 | –0.07 ± 0.5 |
| 30 | 1184/01 | 4 | 0.0–32.5 | 82.2 ± 18.4 | 3.5 ± 2.1 | 40.4 ± 12.8 | 0.23 ± 0.3 | 0.16 ± 0.4 |
| 31 | 1184/02 | 4 | 32.5–60.0 | 79.3 ± 11.3 | 5.5 ± 1.9 | 20.1 ± 11.8 | 0.47 ± 0.4 | 0.42 ± 0.3 |
| 32 | 1184/03 | 4 | 60.0–86.7 | 75.7 ± 12.2 | 5.3 ± 2.0 | 26.5 ± 12.2 | 0.38 ± 0.4 | 0.31 ± 0.3 |
| 33 | 1184/04 | 4 | 86.7–110.8 | 88.5 ± 18.2 | 7.5 ± 2.5 | 28.6 ± 15.4 | –0.13 ± 0.4 | 0.08 ± 0.4 |
| 34 | 1184/05 | 4 | 110.8–133.6 | 97.9 ± 10.7 | 6.7 ± 3.0 | 26.4 ± 15.1 | 0.02 ± 0.4 | 0.05 ± 0.4 |
| 35 | 1184/06 | 4 | 133.6–181.0 | 97.8 ± 17.6 | 7.1 ± 2.8 | 52.2 ± 40.5 | –0.03 ± 0.4 | –0.01 ± 0.4 |
| 36 | 1185/01 | 8 | 0.0–18.5 | 70.5 ± 12.9 | 10.7 ± 2.4 | 18.8 ± 22.4 | 0.17 ± 0.3 | 0.15 ± 0.3 |
| 37 | 1185/02 | 8 | 18.6–29.7 | 71.8 ± 9.5 | 14.4 ± 2.9 | 18.0 ± 4.3 | 0.14 ± 0.3 | 0.04 ± 0.4 |

## C SI 3

Simulation experiment results, including the signed scale coefficient (*m*) used to adjust model solutions for evaluation against data. RMSD values measure the difference between observed perfusion and estimated CBF adjusted by a linear correction with slope *m* (reported here in dimensional form that includes the magnitude of ratio between perfusion and CBF variability). Dagger-flagged PPA in rows indicate data of questionable quality excluded from aggregate analysis.

| $k$ | Expt | PPA | $t_0$ | $m$ | PRx | Mx | PFR |
| --- | --- | --- | --- | --- | --- | --- | --- |
| 1 | 1073/01.08 | 0.0 | 7.5 | +0.48 | 0.64 | 0.71 | p |
| 2 | 1074/01.07 | 0.0 | 9.1 | +23.30 | 0.21 | 0.60 | p |
| 3 | 1074/01.09 | 9.6<sup>†</sup> | 13.5 | +22.84 | 0.03 | 0.71 | p |
| 4 | 1074/01.11 | 7.9<sup>†</sup> | 16.6 | +36.95 | 0.43 | 0.58 | p |
| 5 | 1074/01.12 | 6.7<sup>†</sup> | 18.7 | +14.48 | -0.23 | 0.89 | p |
| 6 | 1074/02.03 | 6.2<sup>†</sup> | 2.4 | +12.49 | 0.23 | 0.48 | p |
| 7 | 1074/02.04 | 3.9 | 4.6 | +3.11 | 0.09 | 0.73 | p |
| 8 | 1074/02.09 | 4.8<sup>†</sup> | 13.2 | +9.59 | 0.07 | 0.78 | p |
| 9 | 1074/02.10 | 2.4 | 15.3 | +5.45 | 0.19 | 0.68 | p |
| 10 | 1074/02.12 | 2.8 | 18.9 | +1.33 | 0.22 | 0.58 | p |
| 11 | 1086/01.07 | 0.0 | 16.1 | +0.27 | 0.35 | 0.86 | z |
| 12 | 1086/01.09 | 0.6 | 20.3 | -0.01 | 0.23 | 0.60 | z |
| 13 | 1086/01.10 | 0.3 | 22.5 | +1.06 | 0.33 | 0.59 | p |
| 14 | 1086/05.04 | 1.1 | 4.6 | +1.39 | 0.56 | 0.72 | p |
| 15 | 1086/05.11 | 0.0 | 19.6 | -0.04 | 0.66 | 0.74 | z |
| 16 | 1086/06.02 | 0.0 | 2.1 | +10.59 | 0.73 | 0.59 | p |
| 17 | 1086/06.03 | 0.0 | 4.2 | -0.43 | 0.49 | 0.43 | z |
| 18 | 1086/06.04 | 0.0 | 6.4 | +2.04 | 0.59 | 0.84 | p |
| 19 | 1086/06.05 | 0.0 | 8.5 | +2.00 | 0.76 | 0.57 | p |
| 20 | 1086/06.06 | 0.0 | 10.6 | +0.81 | 0.61 | 0.63 | p |
| 21 | 1086/06.07 | 0.0 | 12.8 | -0.10 | 0.75 | 0.73 | z |
| 22 | 1086/06.09 | 0.0 | 17.0 | +3.40 | 0.60 | 0.85 | p |
| 23 | 1086/06.11 | 0.0 | 21.3 | +3.72 | 0.50 | 0.58 | p |
| 24 | 1140/01.08 | 0.0 | 11.8 | +0.15 | 0.25 | 0.44 | z |
| 25 | 1141/01.19 | 0.0 | 18.8 | -0.12 | 0.74 | 0.29 | z |
| 26 | 1141/01.72 | 0.0 | 68.3 | +0.05 | 0.82 | 0.26 | z |
| 27 | 1141/01.73 | 0.0 | 70.5 | -0.52 | 0.55 | 0.22 | n |
| 28 | 1141/01.75 | 0.0 | 74.2 | -0.11 | 0.65 | -0.02 | n |
| 29 | 1141/01.76 | 0.0 | 76.4 | +0.67 | 0.64 | 0.30 | p |
| 30 | 1141/01.78 | 0.0 | 80.8 | +5.29 | 0.40 | 0.31 | p |
| 31 | 1141/01.84 | 0.0 | 87.3 | +0.60 | 0.51 | 0.53 | p |
| 32 | 1141/01.93 | 0.0 | 97.9 | +0.10 | 0.20 | 0.25 | z |
| 33 | 1160/01.03 | 0.0 | 7.3 | -2.44 | 0.38 | 0.61 | n |
| 34 | 1165/01.06 | 1.3 | 7.4 | +0.93 | 0.62 | 0.51 | p |
| 35 | 1166/01.12 | 0.0 | 20.0 | +1.52 | 0.16 | 0.60 | p |
| 36 | 1166/01.13 | 0.4 | 22.2 | +1.29 | 0.18 | 0.68 | p |
| 37 | 1166/02.02 | 2.2 | 0.9 | -0.09 | 0.30 | 0.49 | z |
| 38 | 1166/02.03 | 3.4 | 3.0 | -0.02 | 0.41 | 0.51 | z |
| 39 | 1166/02.06 | 3.2 | 9.5 | +6.45 | 0.42 | 0.63 | p |
| 40 | 1166/02.08 | 1.3 | 12.2 | -0.31 | 0.20 | 0.59 | z |
| 41 | 1166/03.03 | 0.0 | 2.5 | +0.08 | 0.20 | 0.51 | z |
| 42 | 1166/03.04 | 0.0 | 4.6 | -0.81 | 0.24 | 0.49 | n |
| 43 | 1166/03.08 | 0.0 | 10.8 | +0.95 | 0.39 | 0.69 | p |
| 44 | 1166/03.10 | 0.0 | 15.1 | +0.24 | 0.18 | 0.61 | p |
| 45 | 1166/03.12 | 0.0 | 19.4 | +0.42 | 0.35 | 0.79 | p |
| 46 | 1166/03.13 | 0.0 | 21.5 | +0.74 | 0.11 | 0.68 | p |
| 47 | 1166/04.02 | 0.0 | 2.1 | -0.61 | 0.05 | 0.33 | z |
| 48 | 1166/04.04 | 0.0 | 6.7 | -0.18 | -0.08 | 0.11 | z |
| 49 | 1166/04.05 | 0.0 | 8.8 | -0.65 | -0.16 | 0.26 | z |
| 50 | 1166/04.06 | 0.0 | 11.0 | -0.52 | -0.22 | 0.16 | n |
| 51 | 1166/04.09 | 0.0 | 17.5 | -0.23 | -0.25 | -0.07 | n |
| 52 | 1183/02.04 | 0.0 | 17.8 | -0.90 | 0.19 | -0.65 | z |
| 53 | 1183/02.07 | 3.0 | 22.9 | -0.94 | 0.24 | -0.53 | n |
| 54 | 1183/02.11 | 3.2 | 30.4 | -3.87 | 0.07 | -0.41 | n |
| 55 | 1183/02.12 | 2.0 | 32.5 | -1.70 | 0.05 | -0.65 | n |
| 56 | 1183/02.16 | 3.7 | 42.3 | -3.69 | 0.01 | -0.70 | n |
| 57 | 1183/02.19 | 1.2 | 49.1 | -1.25 | 0.00 | -0.31 | n |
| 58 | 1183/02.20 | 0.0 | 52.2 | -5.86 | 0.12 | -0.38 | n |
| 59 | 1183/02.24 | 0.6 | 60.1 | -0.25 | 0.07 | -0.65 | z |
| 60 | 1183/03.05 | 1.6 | 10.7 | +2.00 | 0.10 | -0.44 | p |
| 61 | 1183/03.06 | 3.1 | 13.0 | +3.04 | 0.00 | -0.16 | p |
| 62 | 1183/03.08 | 1.9 | 17.1 | +12.39 | -0.05 | -0.31 | p |
| 63 | 1183/03.09 | 4.1<sup>†</sup> | 20.5 | +1.89 | -0.08 | -0.06 | p |
| 64 | 1184/01.09 | 4.0 | 9.2 | -0.24 | 0.07 | 0.18 | n |
| 65 | 1184/01.11 | 0.0 | 13.4 | -0.39 | 0.16 | 0.33 | z |
| 66 | 1184/01.12 | 0.0 | 15.5 | +0.90 | 0.06 | 0.54 | p |
| 67 | 1184/02.02 | 0.7 | 0.7 | -0.03 | 0.13 | 0.63 | z |
| 68 | 1184/02.16 | 0.8 | 20.1 | +11.45 | 0.69 | 0.73 | p |
| 69 | 1184/02.19 | 1.3 | 25.2 | -2.53 | 0.57 | 0.37 | **n** |
| 70 | 1184/03.03 | 2.4 | 4.2 | +7.00 | 0.72 | 0.63 | p |
| 71 | 1184/03.04 | 3.6 | 6.4 | +3.43 | 0.66 | 0.42 | p |
| 72 | 1184/03.06 | 4.3<sup>†</sup> | 10.6 | -6.90 | 0.40 | 0.53 | **n** |
| 73 | 1184/04.03 | 4.0 | 3.8 | -2.73 | -0.25 | 0.28 | **n** |
| 74 | 1184/04.05 | 0.0 | 8.1 | -0.14 | -0.37 | 0.16 | z |
| 75 | 1184/04.09 | 0.0 | 14.4 | -0.37 | -0.14 | 0.13 | **n** |
| 76 | 1184/04.11 | 0.0 | 17.7 | -0.73 | -0.04 | -0.11 | **n** |
| 77 | 1184/05.09 | 0.0 | 12.6 | -0.45 | 0.03 | 0.06 | **n** |
| 78 | 1184/06.01 | 0.0 | 0.0 | -0.48 | 0.40 | -0.09 | **n** |
| 79 | 1184/06.05 | 0.3 | 6.2 | -13.68 | 0.13 | -0.09 | **n** |
| 80 | 1185/01.02 | 0.0 | 1.0 | +0.60 | 0.10 | 0.34 | p |
| 81 | 1185/01.04 | 0.0 | 3.7 | +0.21 | 0.31 | 0.06 | z |
| 82 | 1185/01.05 | 1.2 | 5.9 | +0.08 | 0.29 | 0.07 | z |
| 83 | 1185/01.06 | 0.0 | 8.0 | -0.20 | 0.22 | 0.28 | **n** |

## D SI 4

Table 4: Experiment result summary. Columns tabulate PPA limits, the number of patients represented, the number of represented epochs (folder of patient data), the number of nominal 2-hour experiments conducted, and the corresponding percentage of experiment identified with pPFR (p), zPFR (z), nPFR (n). Experiment results are not influenced by CBF quality measured by PPA values.

Table 4: Experiment result summary.
| | $N_{\text{patients}}$ | $N_{\text{epochs}}$ | $N_{\text{expts}}$ | % PFR (p/z/n) |
| --- | --- | --- | --- | --- |
| All | 11 | 23 | 83 | 46/28/27 |
| PPA≤5 | 11 | 22 | 78 | 44/29/27 |
| PPA≤2 | 11 | 21 | 61 | 39/33/28 |
| PPA≤1 | 10 | 19 | 52 | 38/35/27 |

## E SI 5

**Patient #04-1166** data define several experiments over 4 epochs, who of which are shown in Fig8.

Successful modeling stops around hour 70.15 of their record (between entries 46 and 47) as the dominant regime changes from pPFR to zPFR and nPFR. During pPFR-labeled experiments, the highly-sedate patient suffers autonomic dysregulation characterized by irregular transient ICP elevation after 65 hours. Flow dynamics of this period track the pressure gradient (positive Mx) while perfusion is high (21.5 mL/hg/min) and CPP is ample (75.5 mm Hg), although pressures are low (mean diastolic ABP*≈*55 mm Hg), subcritical ICP*≈*8.5 mm Hg). ABP elevation during the second epoch (lower panel) results in the engagement of CA – thereby decreasing CBF – once the CPP exceeds the lower limit of CA (estimated at about 70 mm Hg.). Model-based experiments fails in these cases because the model lacks the hidden state-dependent (and likely individualized) limits at which autoregulatory engagement occurs or vanishes.

**Patient #04-1183** is featured in an nPFR experiment (Fig4a–b; experiment table entry 53) characterized by a nearly exact negative pressure-flow relationship throughout the interval. The appearance of nPFR here may be explained by volumetric changes resulting from stimuli-instigated tachypnea. Namely, a decrease in CO_2_ initiates vasoconstriction, directly decreasing CBF while indirectly increasing CPP (by lowering ICP) through reduction of vessel-occupied volume. Model estimation fails to account for cardiovascular rate changes and volumetric feedback between vasoregulation and ICP, although MMM data is insufficient at this specific time needed to confirm this (EtCO2 recording is absent at the relevant period).

**Patient #04-1165/01 Experiment 006** is the case featured in the right plot of inconclusive examples. Observed dynamics in this interval are primarily driven by therapeutics, beginning with a decreased of both sedation and vasopressors around 10 minutes, and promptly increased following the hypotensive event. Increased ABP and ICP yield a net increase in CPP, until about 38 minutes into the simulation when mannitol is given to regulate intracranial hypertension. The late, delayed increase in CBF following CPP decrease results from external management rather than endemic processes.

## List of Abbreviations

ABP: arterial blood pressure
CA: cerebral autoregulation
CBF: cerebral blood flow
CPP: cerebral perfusion pressure
CVR: cerebrovascular resistance
EtCO_2_: end-tidal carbon dioxide
EVD: extra-ventricular drainage
GCS: Glasgow Coma Score
GOSES: Glasgow Outcome Scale Extended Score (in figures)
ICP: intracranial pressure
ICHD: intracranial hemodynamics
ICT: intracranial temperature
K-S test: Kolmogorov-Smirnov statistical test
MMM: multimodality monitoring
Mx: mean flow index
P_bt_O_2_: partial pressure of brain tissue oxygen
PFR: pressure-flow relationship
PRx: pressure reactivity (of ICP to ABP) index
PPA: probe placement assistant

